# Impact of a diabetes disease management program on quality of care and costs: propensity score-matched real-world data from Switzerland

**DOI:** 10.1101/2020.07.05.20143438

**Authors:** Maria Carlander, Marc Höglinger, Maria Trottmann, Birgitta Rhomberg, Cornelia Caviglia, Adrian Rohrbasser, Christian Frei, Klaus Eichler

## Abstract

**Objectives:** Structured treatment programs have been recommended for management of patients with chronic conditions to overcome ill-coordinated care. We aimed to evaluate a disease management program (DMP) for diabetes mellitus patients in Switzerland.

**Methods:** We performed a prospective observational study with a propensity score-matched usual care control group from a claims database. We included type-1 and type-2 diabetes patients from a primary care setting. The DMP (intervention) comprised a structured treatment approach with an individual treatment plan, treatment goals and an interprofessional team approach. Our outcome comprehensive measures included quality of life (QOL: EQ-5D-5L), pre-defined indicators for diabetes guideline adherence, number of used services and direct medical costs. We applied a difference-in-difference (DID) approach to compare DMP with usual care (follow-up 1 year). Costs were calculated with non-parametric bootstrapping (2017 Swiss Francs, CHF; conversion rate to Euros: 0.85) from a third-party payer perspective (Swiss health care insurance).

**Results:** QOL in a sub-sample of 80 patients did not change during follow-up (mean utility 0.89 at baseline and follow-up; p=0.94). Guideline adherence showed slight improvements for DMP. For example, “non-adherence” (baseline DMP: 19%) decreased in the DMP group by −3 %-points (DID; 95%-CI: −0.07 to 0.01) but not in the control group. A general trend emerged, though mostly not statistically significant, with less used services in the DMP group compared to the control group. Costs increased in both groups during follow-up, but the increase was higher in the control group (DID, mean total costs per patient per year: CHF −950.00 [95%-CI: −1959.53 to 59.56]). Such a negative difference-in-difference estimate in favor of DMP also emerged for cost sub-categories (e.g. costs for inpatient and outpatient care).

**Conclusions:** The structured treatment program under evaluation is a promising approach to improve diabetes care in a Swiss primary care setting but more follow-up data are needed.

## Background

Structured treatment programs have been recommended for management of patients with chronic conditions, such as diabetes mellitus, to overcome ill-coordinated services across involved providers, strengthen guideline adherence and improve patient outcome. ^1^ Ill-coordinated care can lead to duplication of services and possibly overuse. On the other hand, there is a risk of undertreatment, for example if doctors have too little time for consultations or do not follow evidence-based guidelines. ^2 3^ To meet the high demands for efficient and coordinated care, more and more insurance companies and physician networks are offering structured treatment programs (also known as disease management programs; DMPs) for certain chronic diseases. These evidence-based concepts of care are intended to increase both quality and efficiency of treatment. ^4^

Structured treatment programs for diabetes mellitus have shown mixed results in health care systems similar to Switzerland. A systematic review compiling evidence from 9 evaluations of German DMPs for diabetes patients found that DMPs can improve quality of care. However, some changes in outcome parameters may only be observable over a longer period of time. ^5^ For example, a propensity-score matched controlled study from this review found that the DMP did not reveal any clear medical benefit at even somewhat higher costs. ^4^

In Switzerland, little is known about the impact of structured treatment programs for diabetes mellitus on quality of care and, simultaneously, costs in real world settings. Evaluations are either based on Markov models with data from international literature ^6^, or have separately assessed the impact of diabetes guideline adherence ^7^, financial incentives ^2^ or telemedicine. ^8^ One study with claims data assessed the impact of integrated care (i.e. managed care) for diabetes and found less disease-related hospitalisations and lower costs. ^9^ Recently, an evaluation of diabetes care in a Swiss real-world setting has been published ^10^ and a Swiss cohort study has been set up in the Canton of Valais, but no intermediate results have been communicated yet. ^11^ In summary, the evidence base for diabetes DMPs in Switzerland is scarce. Thus, we evaluated a disease management program with a structured treatment approach for diabetes mellitus patients in Switzerland compared to usual care exploring quality of care, quality of life and costs simultaneously.

## Methods

### Study design

We performed a prospective observational study with a propensity score-matched control group from a claims database.

### Study setting

The study was performed in a Swiss primary care setting (Medbase health care provider). Medbase provides a structured diabetes treatment program within its network of primary care practices since 2017. The first seven practices in the north-eastern part of Switzerland participated in the study and included the diabetes patients. Data were collected on patient and GP group practice level in the years 2017 to 2019. This article presents the mid-term results of the evaluation (1-year follow-up).

In Switzerland, insurees are free to choose their GP in primary care. Structured treatment programs, as described above for the intervention group, may sometimes be applied, but are not implemented on a broader scale in Switzerland. In some contracted insurance models (may be with capitation) physician networks collaborate with insurers and patients joining such networks get rebates on their insurance premiums. The participating practices in our study are serving patients with and without such contracted insurance models.

### Inclusion criteria of patients

We included type-1 and type-2 diabetes patients and all age groups with no gender restriction. Exclusion criteria for the structured treatment program were substantial comorbidities (e.g. dementia), relevant language barriers, living in a nursing home and substantial barriers due to personal living situation or relevant job strains. For all reported outcomes, diabetes patients also had to be insured with SWICA Health Care Organisation, a large Swiss health care insurance (about 1.5 million insurees in 2019).

### Structured treatment program (intervention)

Patients in the intervention group took part in a structured diabetes treatment program. For ease of comparison with other similar programs we use the term DMP within this report.

The DMP under assessment is a structured, evidence-based and patient-centered concept for care of diabetes patients. It comprises an individual treatment plan with treatment goals and an interprofessional team approach. Within the practice network GPs, other physician specialists, physiotherapists, pharmacists and additional health professional are employed. Within the individual treatment plan, specific measures to hit the treatment goals can be adjustment of medication, tailored patient education, periodical control measures and physical training sessions performed by physiotherapists. Current treatment and results of examinations are documented and regularly evaluated together with the patient for achievement of treatment goals and adjustments, if needed. Due to the interdisciplinary team composition, patients are offered a more comprehensive management of their chronic disease compared to a mono-disciplinary approach by physicians. To enhance professional exchange and team competence, regular meetings within professions and within practice teams take place. In addition, quality assurance meetings are held within the physician network, i.e. between practices, to improve and further develop the structured treatment concept.

### Usual diabetes care (control group)

We used a propensity score-matched control group of diabetes patients from the SWICA claims data base (SWICA: about 1.5 million insurees in 2019 in Switzerland; diabetes prevalence in Switzerland: 5% ^1^) to compare the impact of the DMP with secular trends in real-world usual care of diabetes patients. The control patients are treated under the real-world conditions of primary care in Switzerland.

Propensity score matching was performed using the SWICA claims data. We used a propensity score kernel matching approach with “entropy balancing". ^12^ Kernel matching is superior to pairwise matching, because it makes use of all cases in the control group and weights them according to their similarity to the treatment cases. The entropy balancing approach produces a (weighted) control group with means and variances of the matching variables identical to those of the treatment group. Our matching variables were gender, five age-groups; three regions of residence, five categories of urban/rural residence locations; three insurance variables (e.g. high or low deductibles as a reliable indicator for health state); and nine pharmaceutical cost groups (PCG; an indicator for co-morbidities). The matching variables (e.g. PCGs) were extracted for the baseline year 2017 before entering the DMP to exclude contamination by any management decision taken during the DMP itself.

### Outcome measures and data sources

Four main outcome domains of the structured treatment program were evaluated, either on the patient or on the practice level. The outcomes were collected at different points of time (see Figure S1).

#### Quality of Life (patient level)

Self-reported health-related quality of life (QOL) was only assessed in the DMP group with the validated instrument EQ-5D-5L. ^13 14^ The questions include five dimensions of QOL and an evaluation of the overall health status. QOL was collected in a cross-sectional manner in 2018 and 2019. We used several validated translations of the EQ-5D-5L according to the local patient mix (Swiss-German; Italian; French; Spanish; Turkish; Portuguese; Albanian; Serbo-Croatian).

A subsample of patients was selected for assessment of QOL. In each of the seven participating practices, the first 20 consecutive diabetes patients after start of the index period (from March to May, in each year) were asked to fill out the EQ-5D-5L instrument in the group practice prior to consultation. Thus, the information collected via the EQ-5D-5L instrument could be used during the same consultation as monitoring information. After consultation, the QOL data was pseudo-anonymised, linked via an ID number and forwarded to the analysis center.

#### “SGED-Score” (GP group practice level)

We used the SGED-score of the Swiss Society of Diabetology and Endocrinology (SGED). This score was derived to measure good diabetes management in primary healthcare based on international guidelines. ^15^ The criteria of the SGED-score include eight main activities, which should be performed regularly during specific time intervals: controls of clinical and metabolic state, lifestyle advice for overweight and smoking patients, HbA1c measurements, blood pressure measurements, LDL-cholesterol measurements for <75-year-olds, nephropathy test, eye examination and examination of the feet (see Figure S3 for details). For each criterion, a minimum fraction of patients in whom the criteria are to be performed is defined. The SGED score is calculated as a composite indicator (0-100 points) at the practice level for the quality of care of a group of diabetic patients. For example, if the indicator “blood pressure of <140/85 mmHg for at least 65% of diabetes patients” is fulfilled, 15 of 100 points are added to the composite score of the practice. The SGED-score was assed only in the DMP group.

The data needed for the SGED-score were collected by clinicians in the relevant group practices in an Excel-sheet. Data was provided at the practice level, thus no information about blood samples or treatment frequencies in single patients could be drawn from the data sheets.

#### 4 Simple performance measures (4SPM, patient level)

The four simple performance measures (4SPM) indicator provides information about diabetes guideline adherence. This quality of care indicator can be calculated based on patients’ health insurance claims data. ^16^ It comprises information about frequency of 4 performance measures within 1 year (assessments of HbA1c, lipids, nephropathy status and examination at ophthalmologist). Four hierarchical levels of diabetes guideline adherence are defined with the 4SPM:

- Level 0: “non adherent” (no or only one HbA1c-measurements per year)
- Level 1: at least 2 HbA1c-measurements per year
- Level 2: L1 **and** at least 1 measurement of lipids per year
- Level 3: L1, L2 **and** at least 1 control of nephropathy status per year
- Level 4: L1, L2, L3 **and** at least 1 control at ophthalmologist per year

For analysis, each patient can qualify for each level only once, i.e. the share of patients from the 5 levels is summing up to 100%. For two levels, we also applied a modified version:

- Level 3-modified: L1, L2 **and** at least 1 control of nephropathy status per year *or current ACE-inhibitor therapy* (i.e. the drug consequence in case of pathologic nephropathy status)
- Level 4-modified: L1, L2, L3 **and** at least 1 control at ophthalmologist *per 2 years* (extended time interval according to updated Swiss guidelines)

#### Health care services and costs (patient level)

We compared direct medical treatment costs in the structured treatment program (DMP; intervention) with costs of propensity score-matched diabetes patients not participating in the programme (usual care; control).

4SPM, applied services and costs were extracted from the SWICA claims database where diabetes patients who are treated with antidiabetic medication can be identified via diabetes specific PCG-groups. Type 2 diabetes patients without oral drug treatment or insulin cannot be identified and are excluded for these outcome variables. Data were extracted, for example, for demographic attributes (e.g. age, gender, death [yes/no]), health insurance attributes (e.g. managed care coverage [yes/no]), PCG (pharmaceutical cost groups; for diabetes and other morbidity categories), number of services (per person per year), direct medical costs (per person per calendar year, as paid by the health insurance), etc.

### Health economic analysis

Health economic analysis was done from the perspective of a third-party payer (SWICA Health Care Insurance). ^17^ Number of services and direct medical inpatient and outpatient cost data were extracted from the SWICA claims database. Costs included all-diagnosis direct medical costs of all diagnostic procedures, treatments, drugs, complications and consultations covered by the mandatory health insurance company. Data cover the inpatient and outpatient sector for each diabetes patient across all hospitals and all providers.

Inpatient costs in our study represent 45% of the total inpatient costs in Switzerland (a lump sum based on Swiss-DRG tariffs). The remaining 65% of inpatient costs are covered by public authorities in Switzerland and not covered in our analysis. In the outpatient sector, where costs are solely covered by health insurers, each service unit is paid on a fee-for-service base. Thus, direct medical costs in the outpatient sector were calculated via number of reimbursed units (e.g. prescribed medication packages) multiplied with current Swiss basic health insurance tariffs (OKP) per unit. Costs are presented as 2017 Swiss Francs (CHF; official 2017 conversion rate to Euros: 0.85; to US$: 1.02; to British £: 0.76). Out-of-pocket payments and deductibles of the patients were not accounted for. In addition, we did not apply a discount rate for costs due to the short observation period.

### Statistical analysis

For our descriptive analysis, we used means (standard deviation) or medians (interquartile range [IQR]) for continuous variables and proportions for categorical data. For inferential analysis, we applied parametric and non-parametric tests and confidence intervals; p-values <0.05 were considered significant. Confidence intervals of cost data were calculated with non-parametric bootstrapping due to the skew distribution of cost data. ^18^ Analyses were performed using the STATA SE 15 software package (StataCorp. 2015. Stata Statistical Software, College Station, Texas, USA).

## Results

### Patient population

Recruitment of intervention and control group is shown in the study flow (Figure 1) and baseline values are shown in Table 1 (before and after matching). Both groups were well matched with respect to known variables that affect outcomes

**Figure 1:**
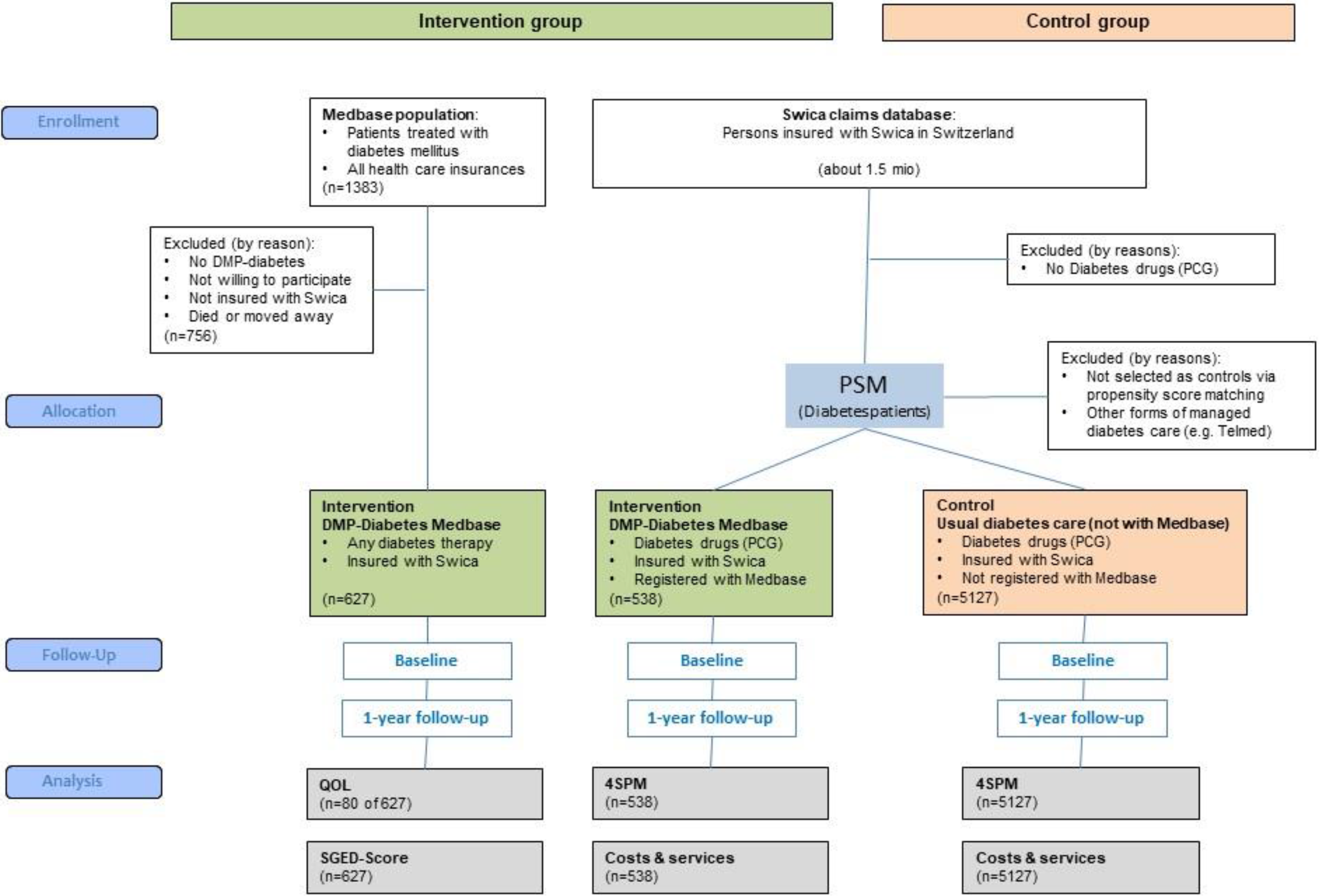
Study flow

**Table 1.**
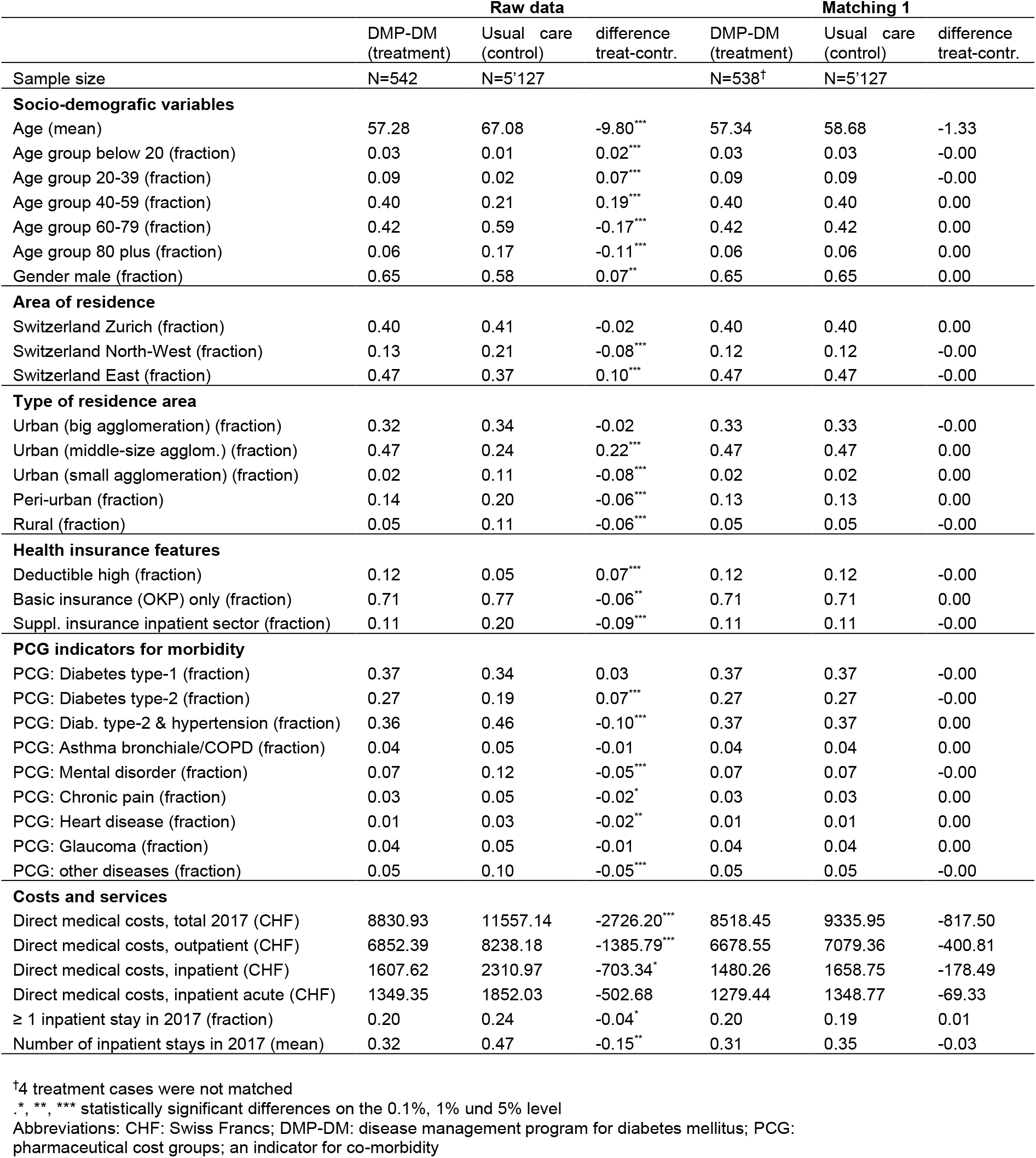
Baseline parameters. Baseline values (2017) of socio-demographic variables, indicators for morbidity, costs and hospitalisations for intervention group and control group (before and after matching).

|  | Raw data |  |  | Matching 1 |  |  |
| --- | --- | --- | --- | --- | --- | --- |
|  | DMP-DM<br>(treatment) | Usual care<br>(control) | difference<br>treat-contr. | DMP-DM<br>(treatment) | Usual care<br>(control) | difference<br>treat-contr. |
| Sample size | N=542 | N=5'127 |  | N=538<sup>†</sup> | N=5'127 |  |
| **Socio-demographic variables** |  |  |  |  |  |  |
| Age (mean) | 57.28 | 67.08 | -9.80*** | 57.34 | 58.68 | -1.33 |
| Age group below 20 (fraction) | 0.03 | 0.01 | 0.02*** | 0.03 | 0.03 | -0.00 |
| Age group 20-39 (fraction) | 0.09 | 0.02 | 0.07*** | 0.09 | 0.09 | -0.00 |
| Age group 40-59 (fraction) | 0.40 | 0.21 | 0.19*** | 0.40 | 0.40 | 0.00 |
| Age group 60-79 (fraction) | 0.42 | 0.59 | -0.17*** | 0.42 | 0.42 | 0.00 |
| Age group 80 plus (fraction) | 0.06 | 0.17 | -0.11*** | 0.06 | 0.06 | 0.00 |
| Gender male (fraction) | 0.65 | 0.58 | 0.07** | 0.65 | 0.65 | 0.00 |
| **Area of residence** |  |  |  |  |  |  |
| Switzerland Zurich (fraction) | 0.40 | 0.41 | -0.02 | 0.40 | 0.40 | 0.00 |
| Switzerland North-West (fraction) | 0.13 | 0.21 | -0.08*** | 0.12 | 0.12 | -0.00 |
| Switzerland East (fraction) | 0.47 | 0.37 | 0.10*** | 0.47 | 0.47 | -0.00 |
| **Type of residence area** |  |  |  |  |  |  |
| Urban (big agglomeration) (fraction) | 0.32 | 0.34 | -0.02 | 0.33 | 0.33 | -0.00 |
| Urban (middle-size agglom.) (fraction) | 0.47 | 0.24 | 0.22*** | 0.47 | 0.47 | 0.00 |
| Urban (small agglomeration) (fraction) | 0.02 | 0.11 | -0.08*** | 0.02 | 0.02 | 0.00 |
| Peri-urban (fraction) | 0.14 | 0.20 | -0.06*** | 0.13 | 0.13 | 0.00 |
| Rural (fraction) | 0.05 | 0.11 | -0.06*** | 0.05 | 0.05 | -0.00 |
| **Health insurance features** |  |  |  |  |  |  |
| Deductible high (fraction) | 0.12 | 0.05 | 0.07*** | 0.12 | 0.12 | -0.00 |
| Basic insurance (OKP) only (fraction) | 0.71 | 0.77 | -0.06** | 0.71 | 0.71 | 0.00 |
| Suppl. insurance inpatient sector (fraction) | 0.11 | 0.20 | -0.09*** | 0.11 | 0.11 | -0.00 |
| **PCG indicators for morbidity** |  |  |  |  |  |  |
| PCG: Diabetes type-1 (fraction) | 0.37 | 0.34 | 0.03 | 0.37 | 0.37 | -0.00 |
| PCG: Diabetes type-2 (fraction) | 0.27 | 0.19 | 0.07*** | 0.27 | 0.27 | -0.00 |
| PCG: Diab. type-2 & hypertension (fraction) | 0.36 | 0.46 | -0.10*** | 0.37 | 0.37 | 0.00 |
| PCG: Asthma bronchiale/COPD (fraction) | 0.04 | 0.05 | -0.01 | 0.04 | 0.04 | 0.00 |
| PCG: Mental disorder (fraction) | 0.07 | 0.12 | -0.05*** | 0.07 | 0.07 | -0.00 |
| PCG: Chronic pain (fraction) | 0.03 | 0.05 | -0.02* | 0.03 | 0.03 | 0.00 |
| PCG: Heart disease (fraction) | 0.01 | 0.03 | -0.02** | 0.01 | 0.01 | 0.00 |
| PCG: Glaucoma (fraction) | 0.04 | 0.05 | -0.01 | 0.04 | 0.04 | 0.00 |
| PCG: other diseases (fraction) | 0.05 | 0.10 | -0.05*** | 0.05 | 0.05 | -0.00 |
| **Costs and services** |  |  |  |  |  |  |
| Direct medical costs, total 2017 (CHF) | 8830.93 | 11557.14 | -2726.20*** | 8518.45 | 9335.95 | -817.50 |
| Direct medical costs, outpatient (CHF) | 6852.39 | 8238.18 | -1385.79*** | 6678.55 | 7079.36 | -400.81 |
| Direct medical costs, inpatient (CHF) | 1607.62 | 2310.97 | -703.34* | 1480.26 | 1658.75 | -178.49 |
| Direct medical costs, inpatient acute (CHF) | 1349.35 | 1852.03 | -502.68 | 1279.44 | 1348.77 | -69.33 |
| ≥ 1 inpatient stay in 2017 (fraction) | 0.20 | 0.24 | -0.04* | 0.20 | 0.19 | 0.01 |
| Number of inpatient stays in 2017 (mean) | 0.32 | 0.47 | -0.15** | 0.31 | 0.35 | -0.03 |

<sup>†</sup>4 treatment cases were not matched
\*, \*\*, \*\*\* statistically significant differences on the 0.1%, 1% und 5% level
Abbreviations: CHF: Swiss Francs; DMP-DM: disease management program for diabetes mellitus; PCG: pharmaceutical cost groups; an indicator for co-morbidity

### Quality of life

Health-related quality of life (EQ-5D) was assessed in a sub-sample of the DMP-DM group in the baseline year 2018 (n=148; thereof 138 patients with valid data) and in 2019 (n=105; thereof 93 patients with valid data). QOL follow-up data was available for 80 patients.

Health-related quality of life (EQ-5D) of the surveyed sub-sample of 80 patients did not change between 2018 and 2019. This was the case for the individual EQ-5D dimensions (Figure 2), the derived utility values (mean utility 0.89 in 2018 and 2019; p=0.94; Figure 3) and for the self-reported health status on the visual analogue scale EQ-5D-VAS (VAS mean in 2018: 77.6; VAS mean in 2019: 79.6; p=0.92; Figure S2). The “pain/discomfort” domain showed the highest share of patients with problems (“slight”, “moderate” “severe”, or “extreme problems”: 2018: 55%; 2019: 57%).

**Figure 2:**
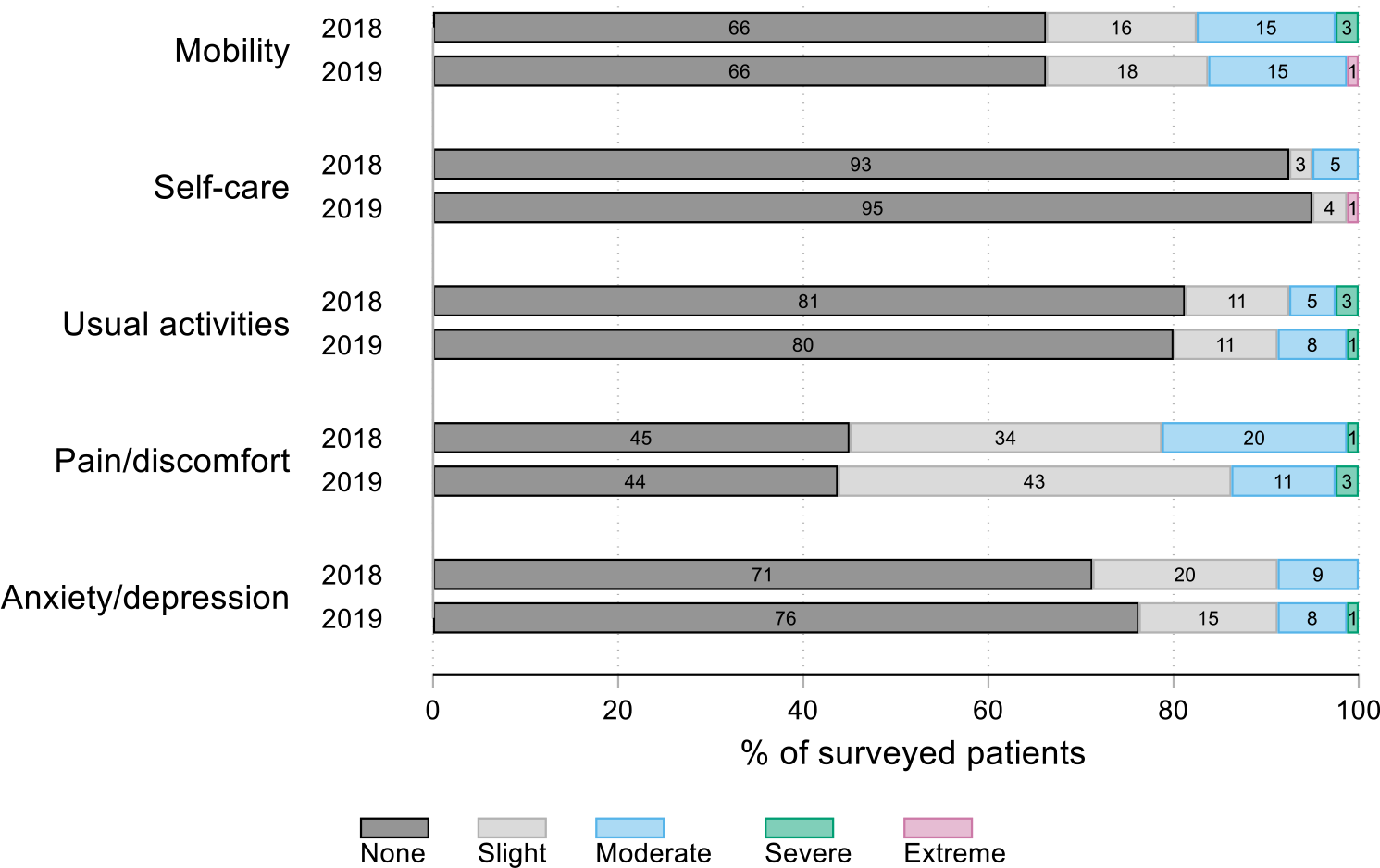
Quality of life (5 dimensions). Share of problems within the 5 QOL dimensions as measured with the EQ-5D-5L descriptive system between 2018 and 2019 (n=80 patients with follow-up data).

**Figure 3:**
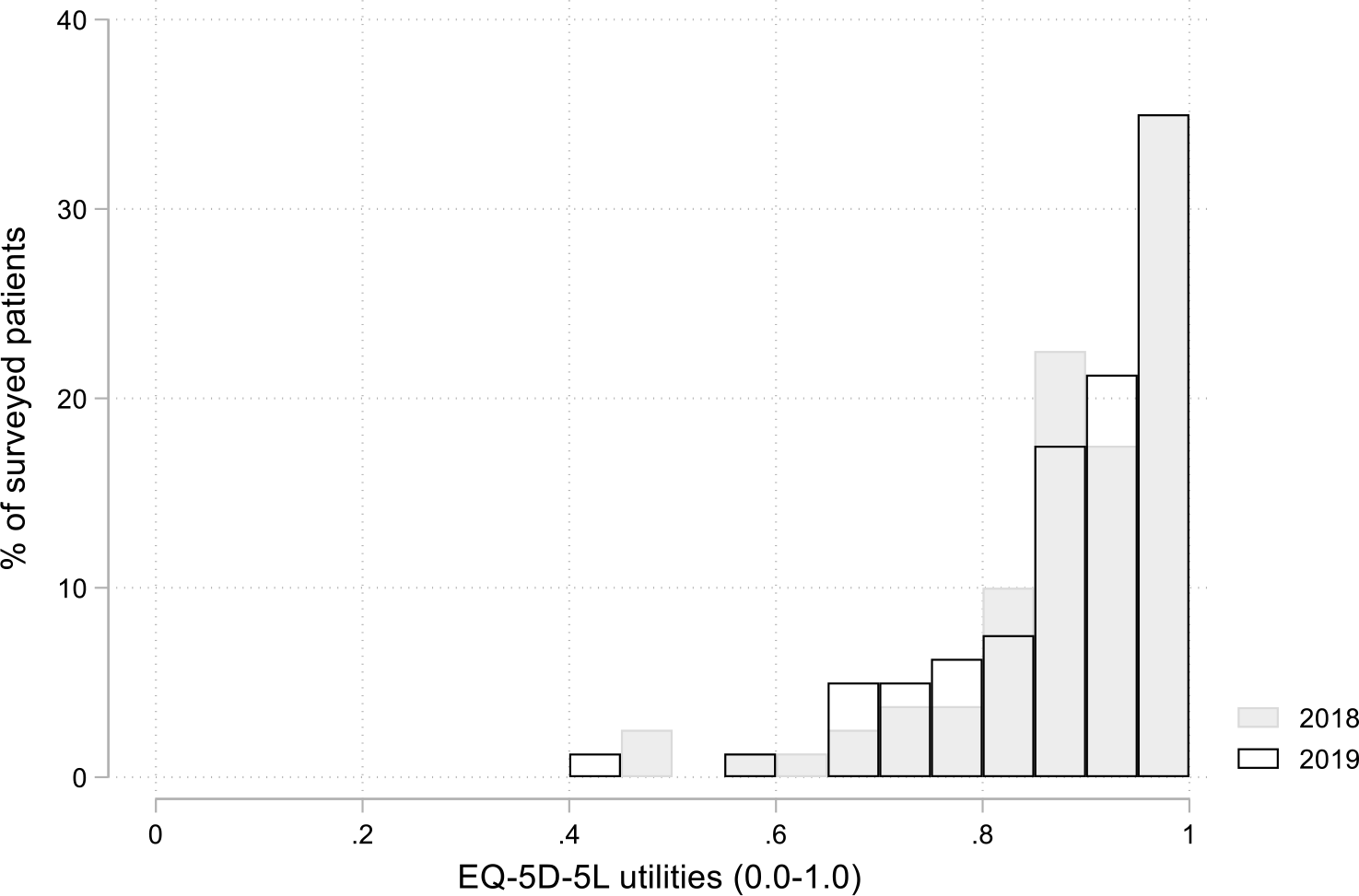
Quality of life (utilities). Health-related quality of life (EQ-5D-5L) as expressed with utility values (range: 0.0-1.0) in 2018 (mean 0.89) and in 2019 (mean 0.89; p=0.94; n=80 patients with follow-up data). The German value set was applied.^14^

### SGED-Score

The SGED score population comprised n=627 diabetes patients insured with SWICA who were treated within the diabetes DMP with and without diabetes drugs. The SGED-Score slightly improved on the practice level from the baseline year 2017 to the follow-up year 2018 (Table 2).

**Table 2.** SGED score overtime (from 2017 to 2018). The SGED score population in 2018 comprised n=627 diabetes patients insured with SWICA who were treated within the structured diabetes treatment program (DMP) with and without diabetes drugs.

| Nr. | Quality criteria | Frequency | Quality indicator | Aim | Maximum points | Number of practices reaching aim 2017 | Number of practices reaching aim 2018 | Range of results across practices (Baseline 2017) N=725 | Range of results across practices (Follow-up 2018) N=627 |
| --- | --- | --- | --- | --- | --- | --- | --- | --- | --- |
| 1 | Regular clinical controls | min. 3 | Diabetes control (single events) | at ≥ 80% of patients | 10 | 0 | 1* | 45.24% – 76.62% | 37.9% – 81.18% |
| 2 | Lifestyle measures | min. 1 | Advice for healthy body weight and physical activity (if BMI ≥ 25)<br>Therapy or advice to stop smoking (for smokers) | at ≥ 80% of patients<br>at ≥ 80% of patients | 5<br>5 | 0<br>2 | 1*<br>2 | 20% – 64.79%<br>28.57% – 85.40% | 20.97% – 83.87%<br>50.0% – 95.54% |
| 3 | HbA1c | min. 2 | HbA1c (DCCT-traceable) | Annual mean:<br><7% at ≥40% of patients<br><8% at ≥60% of patients<br><9% at ≥85% of patients | 5<br>8<br>12 | 1<br>1<br>0 | 3*<br>5*<br>1* | 28.86% – 44.16%<br>42.86% – 62.34%<br>50% – 69.13% | 28.57% – 49.41%<br>51.61% – 72.0%<br>57.26% – 85.6% |
| 4 | Blood pressure | min. 2 | Blood pressure (in sitting position after 5 min. rest) | Annual mean:<br><140/85 mmHg at ≥65% | 15 | 0 | 0 | 8.03% – 33.77% | 5.36% – 33.06% |
| 5 | LDL cholesterol (in patients ≤75 years) | min. 1 | LDL cholesterol in patients ≤75 years (if more than 1 measurement: annual mean) | Annual mean:<br><2.6 mmol/l at ≥63% | 10 | 0 | 0 | 8.62% – 38.66% | 20.88% – 52.87% |
| 6 | Nephropathy assessments | min. 1 | Assessment of serum creatinine + microalbuminuria | at ≥ 80% of patients | 10 | 0 | 0 | 18.75% – 42.86% | 8.06% – 61.18% |
| 7 | Ophtalmologic assessments | min. 1 | Every 2 years ophtalmologic examination by ophtalmologist | at ≥ 80% of patients | 10 | 0 | 0 | 12.99% – 34.13% | 15.29% – 66.07% |
| 8 | Food examinations | min. 1 | (Pulse test + monofilament) OR (pulse test + vibration sense) | at ≥ 80% of patients | 10 | 0 | 0 | 3.75% – 35.71% | 9.68% – 64.8% |

\*Improvement in number of practices reaching aim in 2018;
Abbreviations: DMP: disease management program (for diabetes mellitus);

In 2017, before implementing the DMP, three of 11 SGED criteria were fulfilled in at least one of the 7 participating group practices. In 2018, after 1 year of follow-up, six of 11 SGED criteria were fulfilled in at least one of the 7 participating group practices. The most often fulfilled criterion (in 5 of 7 practices) was “at least 2 HbA1c measurements per year in ≥ 60% of diabetes patients with a HbA1c level of < 8%”. The number of SGED criteria that was not fulfilled in any practice decreased from 8 of 11 criteria in 2017 to 5 of 11 criteria in 2018.

### 4 simple performance measures (4SPM)

The 4SPM population comprised the matched study population (n=538 diabetes patients in the DMP group; n=5127 diabetes patients in the control group). We found slight improvements in the 4SPM indicators in the DMP group compared to the control group in our difference-in-difference analysis (Baseline: 2017; follow-up: 2018).

At baseline, management of 19%-20% of patients was classified as “non-adherence” according to the pre-defined criteria. 25% of patients qualified for Level 1, 37%-40% for Level 2, 3% to 5% for Level 3 and 11% to 13% for Level 4 (Table 3). During follow-up, the share of patients in two lower adherence levels decreased in the DMP group compared to usual care (DID: for “non-adherence” Level 0: −3%-points; Level 2: −4%-points). For Level 4 (adapted criteria: Level 3 plus one ophthalmologic assessment per 2 years), a significant difference-in-difference improvement for DMP emerged with an increase of 7%-points (95%-CI: 3% to 12%).

**Table 3.**
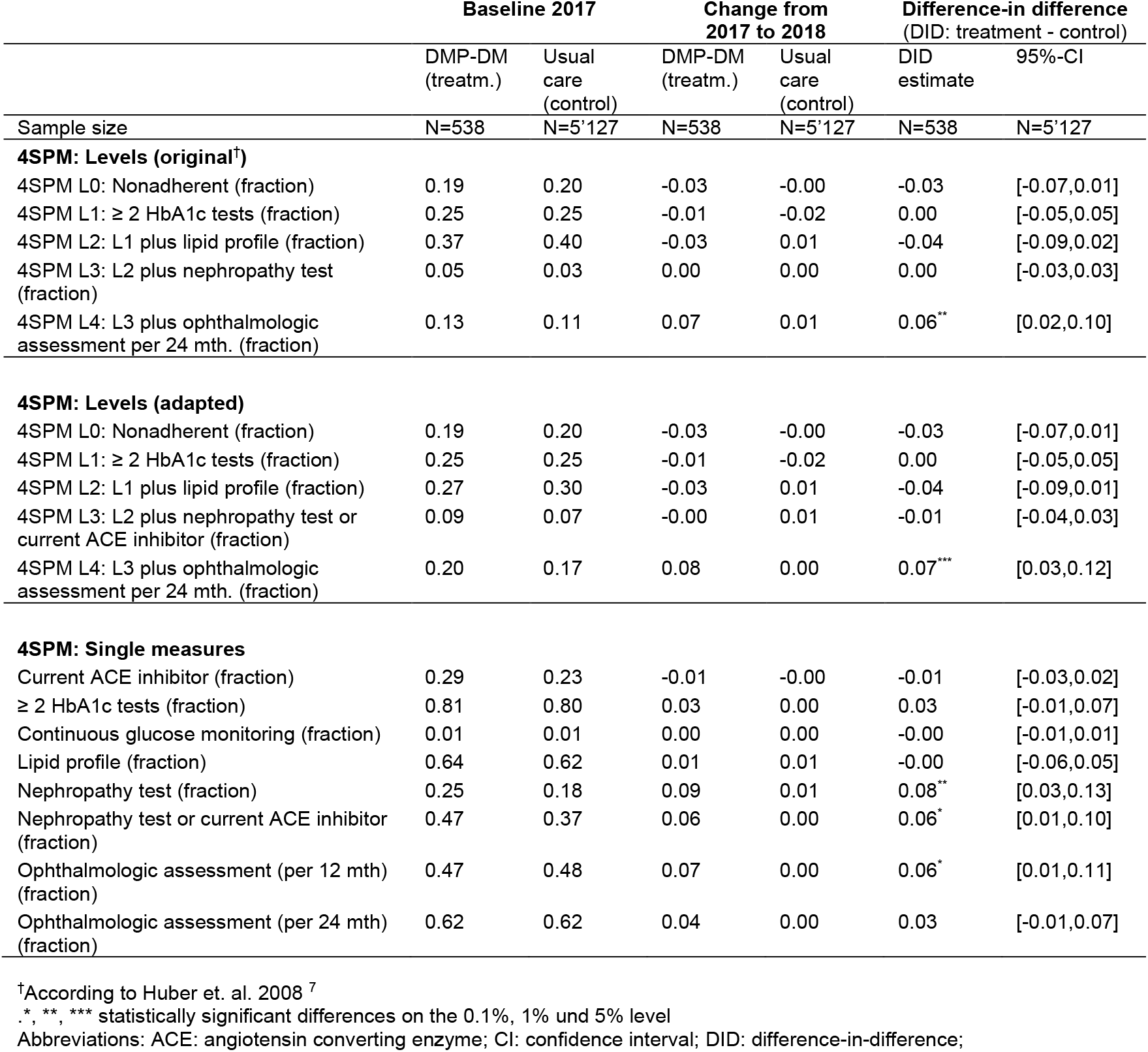
Four Simple Performance Measures (4SPM) over time. Baseline values (2017), change from 2017 to 2018 and difference-in-difference (DID) estimates are depicted.

Analysis of single performance measures (i.e. not the Level view) showed, that improvements in the DMP group were seen for the adherence to nephropathy related guidance (plus 8% of patients [95%-CI: 3% to 13%]) and guidance related to ophthalmologic examinations (plus 6% of patients [95%-CI: 1% to 11%]).

### Health care services and costs

The population for analysis of health care services and costs comprised the matched study population (n=538 diabetes patients in the DMP group; n=5127 diabetes patients in the control group).

In our difference-in-difference analysis (Baseline: 2017; follow-up: 2018), a general trend emerged, though mostly not statistically significant, with less applied services and lower costs in the DMP group compared to the control group. For example, baseline total direct medical costs (comprising inpatient and outpatient sector treatment) in our matched sample were high in both groups with between CHF 8520 (DMP group) and CHF 9336 (control group) per year. These costs increased in both groups during follow-up, but the increase was about six times higher in the control group. This led to a mean cost difference-in-difference (DID) of CHF −950.00 (95%-CI: −1959.53 to 59.56) per patient per year in the DMP group compared to the control group (Table 4). Such a negative difference-in-difference estimate in favor of DMP also showed up with cost sub-categories, such as cost for inpatient and outpatient treatment separately. Furthermore, the fraction of diabetes patients with at least one inpatient treatment per year during follow-up decreased significantly by −6% (95%-CI:-11% to −1%) in the DMP group (baseline rates in 2017: intervention group 20%; control group 19%).

**Table 4.** Costs and medical services over time. Baseline values (2017), change from 2017 to 2018 and difference-in-difference (DID) estimates are depicted. Costs are direct medical costs, applied services are from the inpatient or outpatient sector.

|  | Baseline 2017 |  | Change from 2017 to 2018 |  | Difference-in difference (DID: treatment - control) |  |
| --- | --- | --- | --- | --- | --- | --- |
|  | DMP-DM (treatment) | Usual care (control) | DMP-DM (treatment) | Usual care (control) | DID estimate | 95%-CI |
| Sample size | N=538 | N=5127 | N=538 | N=5127 | N=538 | N=5127 |
| **Costs (main categories): CHF** |  |  |  |  |  |  |
| Treatment costs (total) | 8518.45 | 9335.95 | 170.65 | 1120.64 | -949.98 | [-1959.53,59.56] |
| Treatment costs, outpatient<sup>†</sup> | 6678.55 | 7079.36 | -12.04 | 276.84 | -288.87 | [-770.88,193.13] |
| Treatment costs, all inpatient | 1480.26 | 1658.75 | 137.50 | 485.31 | -347.81 | [-1117.32,421.70] |
| Treatment costs, acute care inpatient | 1279.44 | 1348.77 | 173.80 | 556.98 | -383.18 | [-998.19,231.84] |
| **Costs (outpatient sector<sup>†</sup>): CHF** |  |  |  |  |  |  |
| Physician consultations | 1212.39 | 1173.17 | -36.29 | -6.38 | -29.91 | [-171.48,111.66] |
| Physiotherapy | 243.44 | 227.49 | -7.80 | 26.29 | -34.09 | [-86.41,18.24] |
| Occupational therapy | 18.38 | 22.96 | 4.96 | 0.16 | 4.80 | [-21.56,31.15] |
| Psychiatric care | 148.00 | 163.21 | -3.49 | 18.76 | -22.25 | [-73.19,28.69] |
| Therapy at psychologist | 0.00 | 0.00 | 0.00 | 0.00 | 0.00 | [0.00,0.00] |
| Medication KVG | 2262.39 | 2564.13 | -96.77 | 44.42 | -141.19 | [-402.53,120.15] |
| Hospital care, outpatient | 745.08 | 741.14 | 36.50 | -12.92 | 49.42 | [-158.58,257.42] |
| Blood tests and other analyses | 452.38 | 488.48 | 39.77 | 40.25 | -0.49 | [-51.62,50.64] |
| X-ray examinations | 317.60 | 337.32 | -12.78 | 3.97 | -16.75 | [-84.28,50.78] |
| Nutrition advice KVG | 26.57 | 15.80 | 2.62 | 1.49 | 1.13 | [-8.41,10.66] |
| Diabetes nurse KVG | 28.66 | 19.20 | -2.68 | -3.32 | 0.64 | [-8.75,10.03] |
| **Medical Services** |  |  |  |  |  |  |
| ≥ 1 inpatient stay (fraction) | 0.20 | 0.19 | -0.04 | 0.02 | -0.06<sup>*</sup> | [-0.11,-0.01] |
| Number of inpatient stays (mean) | 0.31 | 0.35 | -0.03 | 0.05 | -0.08 | [-0.19,0.03] |
| Number of outpatient consultations (mean) | 9.69 | 10.34 | -0.11 | 0.60 | -0.71<sup>*</sup> | [-1.41,-0.01] |
| Consultation at diabetes specialist (fraction) | 0.25 | 0.25 | 0.04 | 0.03 | 0.02 | [-0.02,0.05] |
| >1 consultation at diabetes specialist (fraction) | 0.20 | 0.21 | 0.04 | 0.03 | 0.01 | [-0.02,0.05] |
| Hospital care, outpatient (CHF 500+) (fraction) | 0.31 | 0.32 | 0.02 | 0.02 | -0.00 | [-0.05,0.05] |

<sup>†</sup> Outpatient costs do not add up to total outpatient costs as only main outpatient cost categories are shown;
\*, \*\*, \*\*\* statistically significant differences on the 0.1%, 1% und 5% level
Abbreviations: CHF: Swiss Francs; CI: confidence interval; DID: difference-in-difference; KVG: (Swiss) Krankenversicherungs-Gesetz (Swiss Basic Health Insurance Act);

## Discussion

After one year of follow-up, the diabetes disease management program under investigation was associated with improvements in quality of care, stable QOL and lower costs compared to the propensity score matched control group in our difference-in-difference analysis.

### Strengths and limitations

Our study has several strengths. We report real-world data of a big Swiss health insurance organisation which adds evidence about the impact of a structured diabetes care approach in primary care of a social health insurance system. Assessing patient benefit and costs simultaneously is an important health services research issue. With this approach, the value of health care for specific chronic conditions can be better understood. ^19^

Our study has, however, several limitations. First, this is an observational study and causal inference can, strictly speaking, not be drawn. However, we addressed the problem of confounding with a thorough propensity score matching approach generating a control group from a big Swiss claims data base. Second, we have no detailed clinical data on the patient level to further describe our diabetes population (e.g. type of diabetes; drug therapy). However, we report validated indicators for morbidity as represented via PCGs. ^20 21^ Third, diabetes patients without specific diabetes medication are not included for the outcomes 4SPM, costs and services because, in Switzerland, health care insurances are not allowed to hold specific diagnoses of their patients within the claims data base. Fourth, the follow-up of our study is short. A longer follow-up is needed to confirm or refute our results. Variability in costs is high, even in patients with chronic conditions with multiple service use. Thus, “regression to the mean” may lead to non-significant cost differences in the opposite direction during longer follow-up. Fifth, QOL was measured only in a small sub-sample of patients and we do not have specific clinical data to describe this population in detail. However, even though a random sample was not possible due to logistic problems in the practices, we believe that our sampling process of consecutive patients within the defined sampling period has led to a reasonably representative selection from our total diabetes population. Nevertheless, results should to be interpreted cautiously and strong conclusions may be drawn from our QOL results. Finally, the data quality for the SGED score items may have limitations, as practices have first initiated routine data collection for these indicators recently. Peer discussion and quality rounds are installed now to further increase learning from study results.

### Comparison with other studies

QOL was unchanged in our study which is in line with other trials. A cluster randomized trial from Austria tested a DMP for type 2 diabetes patients and found no significant differences in EQ-5D-3L VAS compared to usual care after one-year follow-up. ^22 23^ Baseline QOL was similar to our sample (EQ-Index 0.88; VAS: 71.1), as also confirmed in a cross-sectional study of diabetes patients from Canada with an EQ-5D index of 0.85. ^24^ A German cross-sectional study found slight improvements of QOL in patients joining a diabetes DMP (reported range of EQ-index in seven classes of co-morbidities: DMP 0.83-0.54; usual care: 0.70-0.40) ^25^ In summary, these comparisons provide indirect evidence that our sampling procedure for QOL measurement may have led to no substantial selection bias.

For the SGED score only one published assessment was found. In a sample of Swiss urban group practices (Sanacare) the SGED score was assessed in 235 diabetes patients who were treated according to the chronic care management model. ^10^ The authors report high guideline adherence fulfilling all but one SGED criteria in their sample (not fulfilled: “≥ 65% of patients with a blood pressure of < 140/90 mmHg”). Observed differences to our results with seemingly lower adherence may be due to poor documentation of clinical procedures and testing, low data quality or, indeed, different compliance with current guidelines.

A reasonably high concordance of our 4SPM results with data from another Swiss claims database ^7^ supports reliability of this indicator. Non-adherence (i.e. Level 0) is somewhat less frequent in our sample (20% vs. 30%), while Level 1 (25% vs. 24%), Level 2 (37% vs. 37%) and Level 3 (4% vs. 4%) show nearly identical frequency. Applying the new Swiss guidance for one ophthalmologic control within *two* years translates into higher adherence in our sample compared to the older published data (13% vs. 5%).

Comparison of cost and service data across health care systems are problematic due to different service structures and incentives. ^17^ Another big health care insurance in Switzerland reports mean annual direct medical costs of diabetes patients being in a range between CHF 2’910 and CHF 12’930 (costing year 2011), depending on the number of co-morbidities ^20^, and between CHF 9’466 and CHF 10’530 (costing years 2012-2013), depending on the integration of services. ^9^ Thus, mean annual costs in our sample between CHF 8’830 and CHF 11’557 (costing year 2017; raw data) are in a similar range.

### Implications for clinicians and public health decision makers

Our study indicates that implementing a structured treatment program for diabetes patients in primary care is a promising option. This has implications for clinicians and managers of health care organisations alike.

Monitoring of quality of care for chronic conditions is of high importance to judge if patients are treated according to current standards. The SGED score, as used in our study, is a promising tool but needs further testing in the field. For example, more data collection is needed from diverse providers to arrive at a representative benchmarking data pool. In addition, minimal important differences for change should be defined. Further measurement cycles are needed to better understand the data.

Measuring QOL during practice consultations offers a unique opportunity to involve patients in the treatment process and get a better understanding of disease progression from the patients’ viewpoint. ^19 26^ Furthermore, comparing patient benefit with treatment costs is a cornerstone in judging the value of health care. ^19^ Leading international hospitals have recently engaged in implementing QOL measurements in their clinical routine procedures in the inpatient and outpatient sector to show excellence. ^27^ Also, some Swiss public authorities have made the assessment of patient-reported outcomes a pre-requisite for public funding in orthopedic surgery. ^28^ The assessment of PROMs may soon be a standard of care for patients with chronic conditions and diabetes care would be a worthful testing field as shown in our physician practice network.

The current magnitude of savings of about 10% of annual costs of diabetes patients is relevant from the payers’ perspective. However, cost differences are not statistically significant and follow-up is short. If these results can be confirmed after longer follow-up, such structured treatment programs are an example of value-based health care, as similar or better quality of care can be provided at lower cost.

## Conclusions

The structured treatment program under evaluation is a promising approach to improve diabetes care in a Swiss primary care setting, but more follow-up data are needed.

## Data Availability

The health insurance claims data as used for our analysis are stored at Swica Health Organisation, Switzerland. Any questions concerning these data should be directed to MT.
Data for QOL and SGED score are stored at Winterthur Institute of Health Economics. Any questions concerning these data should be directed to MC.

## Glossary

CG: Control group
CHF: Swiss francs
CI: Confidence intervall
DM: Diabetes mellitus
DMP: Disease Management Program
IG: Intervention group
OKP: Obligatorische Kranken-und Pflegeversicherung (Basic health insurance in Switzerland)
PCG: Pharmaceutical cost groups
PROM: Patient-reported outcome measure
QOL: Quality of life
SD: Standard deviation
SGED: Schweizerische Gesellschaft für Endokrinologie und Diabetes (Swiss Society for Endocrinology and Diabetes)
VAS: Visual analogue scale
4SPM: 4 simple performance measures (HbA1c; lipid profile; nephropathy examination; ophthalmologic examination)

## Declarations

### Ethics approval and consent to participate

Not applicable. According to the local Ethics committee (BASEC-Nr. Req-2017-00416), patient consent was not necessary as our evaluation using an anonymised database does not fall under the Human Research Act in Switzerland.

### Competing interests

KE, MC, MH report grants from SWICA Health Care Organisation, Switzerland, during the conduct of the study; KE and MH also report grants from Swiss Accident Insurance (SUVA), grants from AO Foundation, grants from Swiss Federal Office of Public Health, outside the submitted work; KE also reports grants from Astellas, grants from AstraZeneca, grants from Celgene, grants from Janssen-Cilag, grants from Novartis, grants from Amgen, outside the submitted work; BR, MT, CF are employed by SWICA Health Care Organisation, Switzerland; CC and AR are employed by Medbase health care provider, Switzerland; AR is also employed by Institute of Primary Health Care (BIHAM), University of Bern, Switzerland; SWICA Health Care Organization holds shares of Medbase health care provider.

### Funding

The study was in part financially supported by SWICA Health Care Organization, Switzerland. Funding was paid to the Winterthur Institute of Health Economics Institute at Zurich University of Applied Sciences. The funding body commented on the final draft of the manuscript but did not make final decisions regarding the design of the study, the data collection, the analysis and the interpretation of results.

### Authors’ Contributions

KE is the guarantor and drafted the manuscript. All authors contributed to the development of the selection criteria and data extraction criteria. MT managed the claims database and extracted data. AR and CC managed the Medbase database. MH, MC, AR and MT provided statistical expertise and performed the analysis. All authors contributed to interpretation of results. All authors read, provided feedback and approved the final manuscript.

## Acknowledgements

The authors are grateful for the valuable support of all diabetes patients joining the structured treatment program and providing QOL data; furthermore, our thank goes to all involved staff from all health professions within the participating Medbase group practices. Without their engagement this study would not have been possible in the present form.

Participating Medbase practices (in alphabetical order of location):

- Medbase Basel
- Medbase Eglisau, Zurich
- Medbase Oerlikon, Zurich
- Medbase St.Gallen
- Medbase Wiedikon, Zurich
- Medbase Wil, St.Gallen
- Medbase Winterthur, Zurich

## Supplement

**Figure S1:**
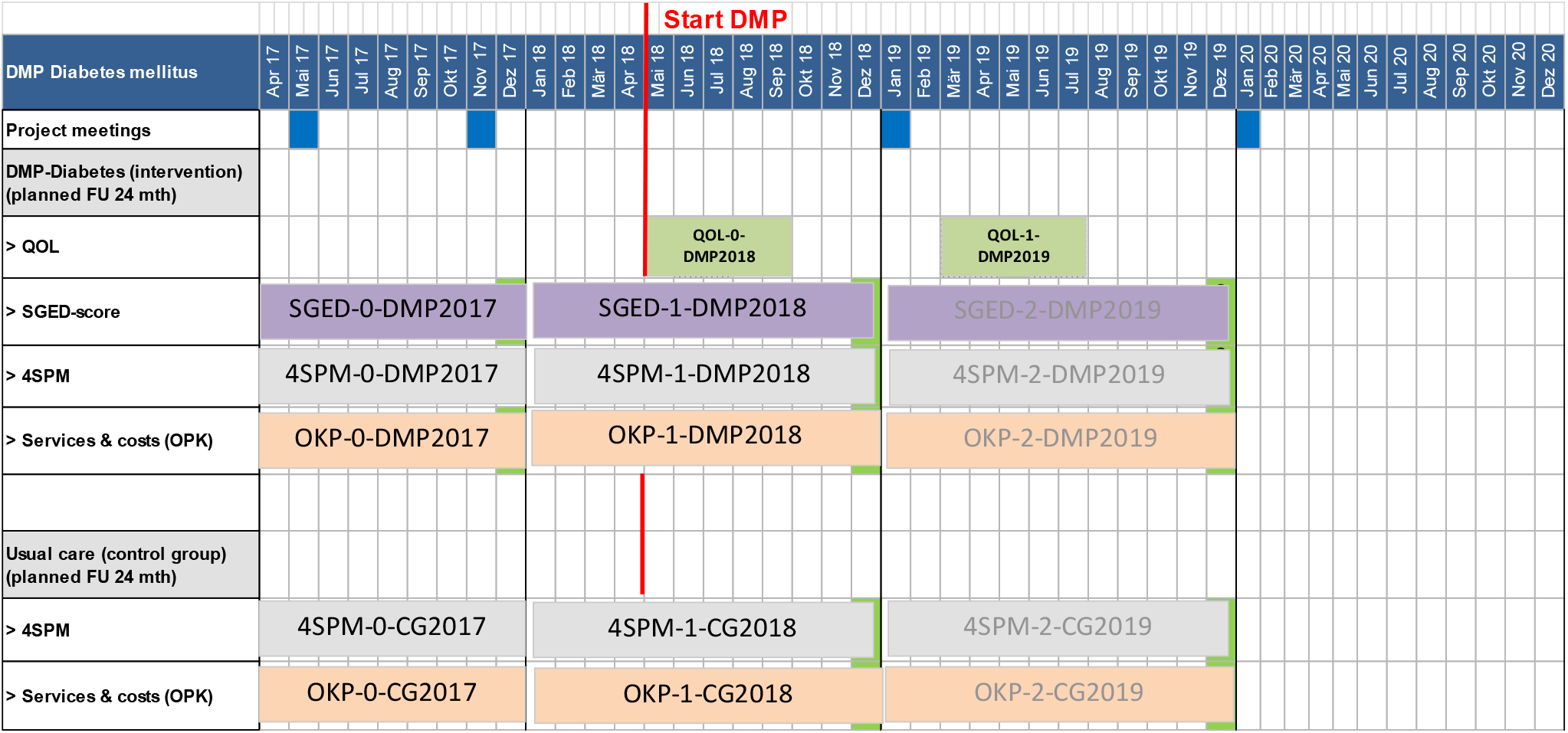
Data collection over time. Time windows of data collection are shown for the intervention group (DMP-Diabetes) and the control group (usual care) for each of the outcome measures. Projected data analysis for SGED, 4SPM and OKP measures for time window 2019 is also shown but will not be available before end of 2020. **Abbreviations**: DMP: disease management program; FU: follow-up; QOL: quality of life; SGED: Schweizerische Gesellschaft für Endokrinologie und Diabetes; 4SPM: Four simple performance measures; OKP: obligatorische Kranken-und Pflegeversicherung (Swiss basic health insurance);

**Figure S2:**
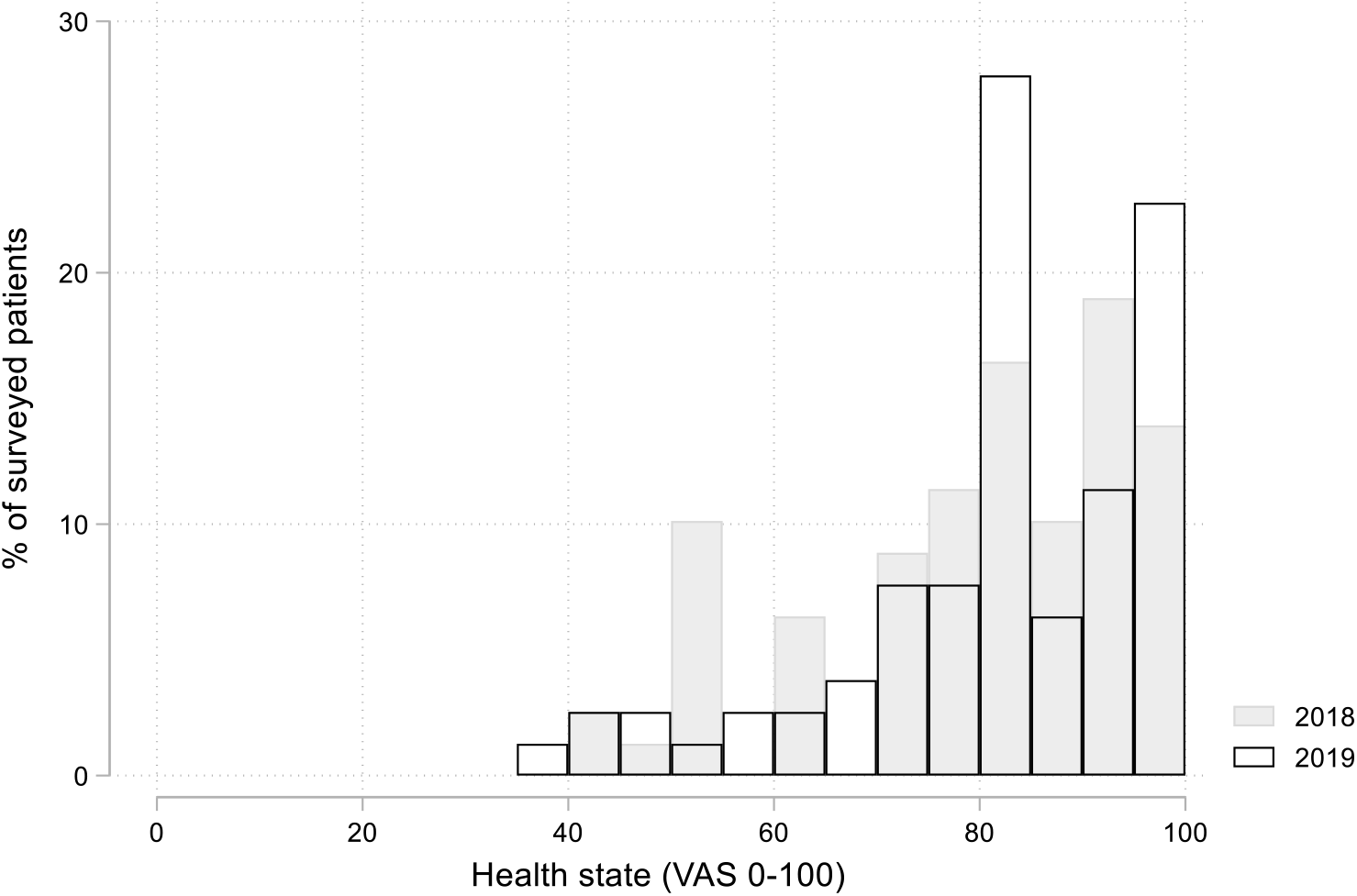
Health state (VAS). Self-estimated health state (EQ-5D-5L) as expressed with visual analogue scale (VAS: range: 0-100) in 2018 (mean 77.6) and in 2019 (mean 79.6; p=0.92).

**Figure S3:**
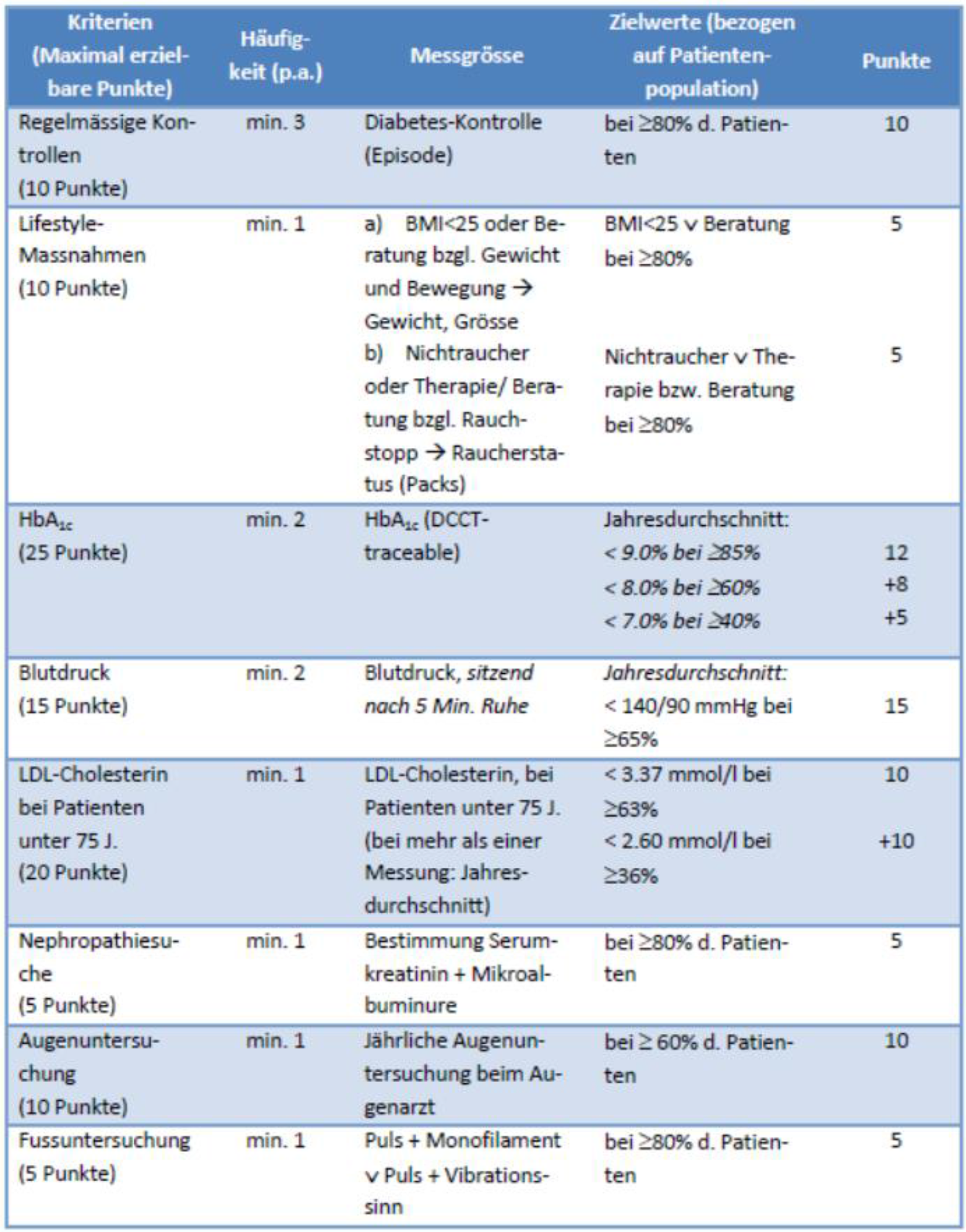
SGED Score criteria. ^15^

## Notes

### Clinical Trial

The study was not registered as we compared observational data with claims data.

### Author Declarations

Ethical approval was not required according to the local Ethics committee (decision of Kantonale Ethikkommission Zurich, Switzerland, on our formal request: BASEC-Nr. Req-2017-00416). Our evaluation does not fall under the Human Research Act in Switzerland as it is solely a quality control evaluation for institution-internal purposes.

